# Neural network pattern recognition of ultrasound image gray scale intensity histogram of breast lesions to differentiate between benign and malignant lesions

**DOI:** 10.1101/2020.05.01.20088245

**Authors:** Arivan Ramachandran, KR Shiva Balan, Swathi Kiran, Mohamed Azharudeen

## Abstract

The aim of this study is to analyze the effectiveness of grayscale intensity histogram to differentiate benign and malignant lesions using a convolutional neural network. Data (200 USG images, 100-malignant, 100-benign) was downloaded from an online access repository. The images were despeckled using ImageJ software and the grayscale intensity histogram values were extracted. In-built neural network pattern recognition application in Matlab R2019b was used to classify the images, which is a two-layer feed-forward network, with sigmoid hidden and softmax output neurons. The positive predictive value of the CNN was 95%. The best performance of 0.078264 was achieved at 36 epochs in the validation set. This study suggests that the grayscale intensity histogram of a USG image is an easy and feasible method to identify malignant lesions through an artificial neural network.

## INTRODUCTION

Breast cancer is the most common cancer in Indian women with a prevalence of 25.8 per 1,00,000. Lack of adequate breast cancer screening, diagnosis at a later stage, and unavailability of resources are quoted as the main reasons for the increase in mortality in breast cancer patients in India (1). Multiple imaging modalities like USG, X-ray mammography, CT, PET, and MRI are being used to screen, diagnose, and evaluate breast cancer.

Ultrasound is the imaging modality of choice in suspicious breast lesions in young women and in pregnant women. Ultrasound has higher accuracy and sensitivity in the detection of malignant lesions compared to X-ray mammography (2).

Usage of Artificial Intelligence (AI) for image recognition and classification is an upcoming method and can be implemented in areas with resource and man-power limitations as it is suggested that neural network-based differentiation of breast lesions has the capacity to significantly reduce unnecessary biopsies and can perform equivalent to trained human radiologists (3,4). In this study, we are evaluating the efficiency of Convolutional Neural Network (CNN) in classifying malignant and benign breast USG images downloaded from an online dataset based on their grayscale intensity histograms.

## MATERIALS

- 200 USG Images (100 malignant and 100 benign) downloaded from https://data.mendeley.com/datasets/wmy84gzngw/1
- Matlab R2019b software
- ImageJ software

VARIABLE: Gray-scale intensity histogram

## METHODOLOGY

Ultrasound images of 100 malignant and 100 benign breast lesions were randomly downloaded from an open access repository https://data.mendeley.com/datasets/wmy84gzngw/1(5). The images were then loaded in the ImageJ software and despeckling was done to improve the contrast resolution of the images (6).

Region of interest (ROI) was drawn over the breast lesions in all the 200 images by a board-certified radiologist and the gray-scale intensity histogram values were extracted. The values were entered in a datasheet and were imported to the Matlab R2019b software.

A total of 200 histograms were divided into

1. Training set containing 70% of total images (140 out of 200)
2. Validation set containing 15% of total images (30 out of 200)
3. Test set containing 15% of total images (30 out of 200)

The in-built application of Matlab R2018b Neural net recognition was used. It is a two-layer feed-forward network, with sigmoid hidden and softmax output neurons. The network was trained with scaled conjugate backpropagation. In our study, we used 30 hidden neurons (7). An input file was created with 200 histogram values as a 255×200 matrix datasheet. The target output was assigned as 1 for malignant and 0 for benign using a 2×200 matrix datasheet. The software trained the CNN by a randomized selection of the inputs automatically.

## RESULTS

The automated results were obtained, and the following figures were created.

The prediction percentages were recorded in a confusion matrix. The positive prediction in the training set, validation set, test set and the combined set is 95.7%, 96.7%, 90% and 95% respectively (Figure. 1).

**FIGURE. 1.**
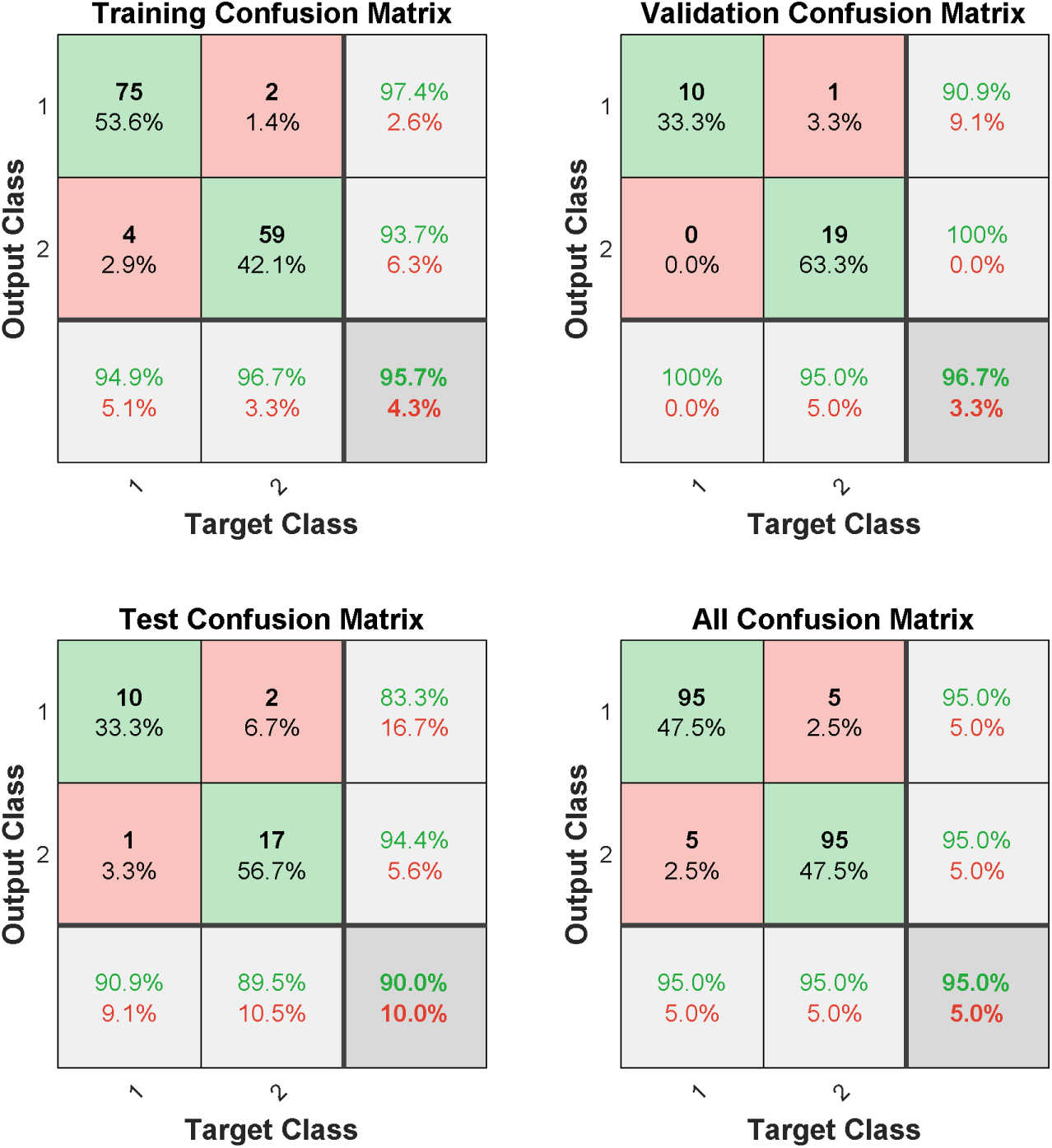
Confusion matrices of training, validation, test and combined sets

The best performance of 0.078264 was achieved at 36 epochs in the validation set after which the cross-entropy increased, suggesting an increase in training errors (Figure. 2)

**FIGURE. 2.**
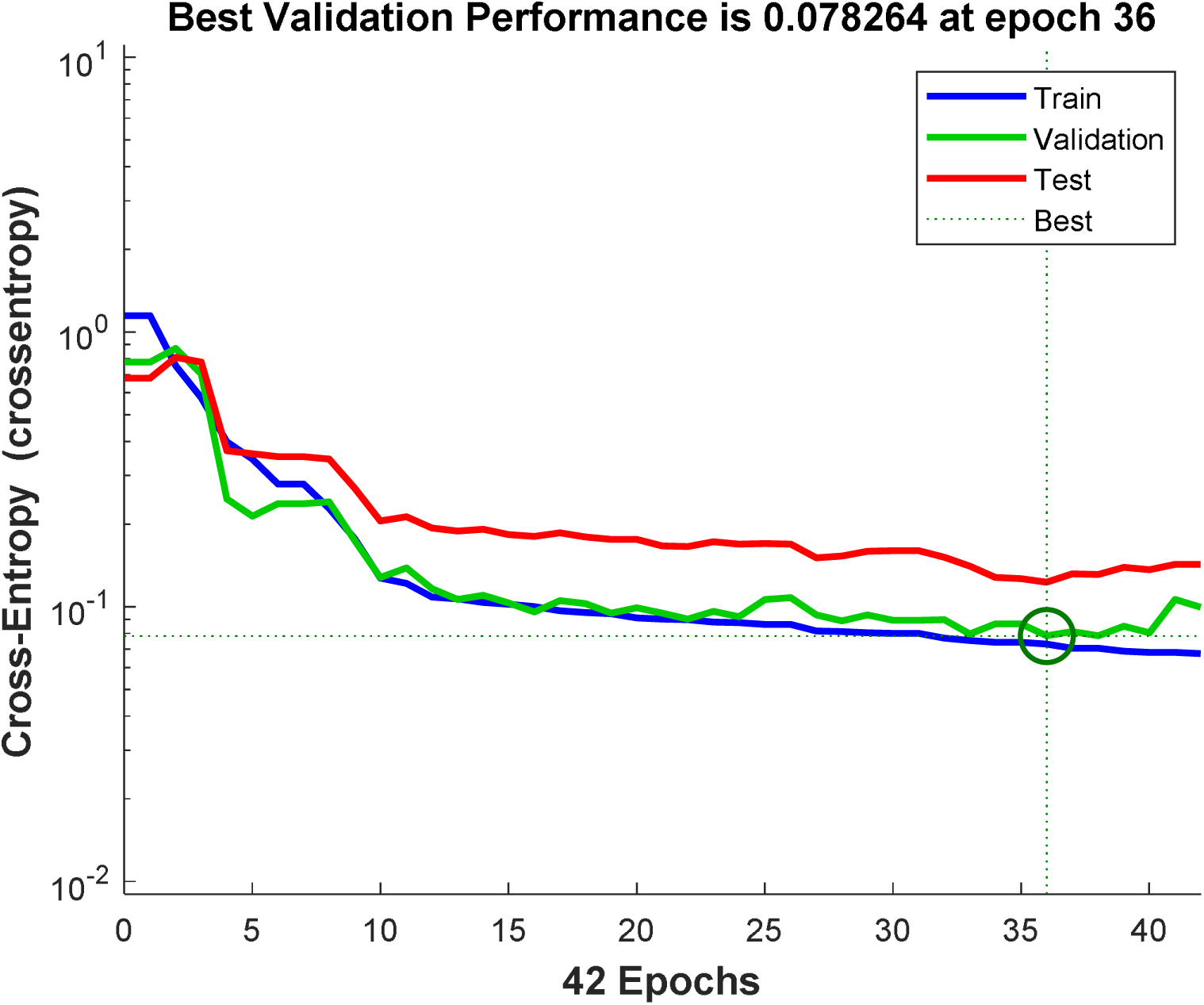
Performance plot of Matlab2019b CNN

Based on the ROC curves (Figure. 3), the positive predictive value was 95%.

**FIGURE. 3.**
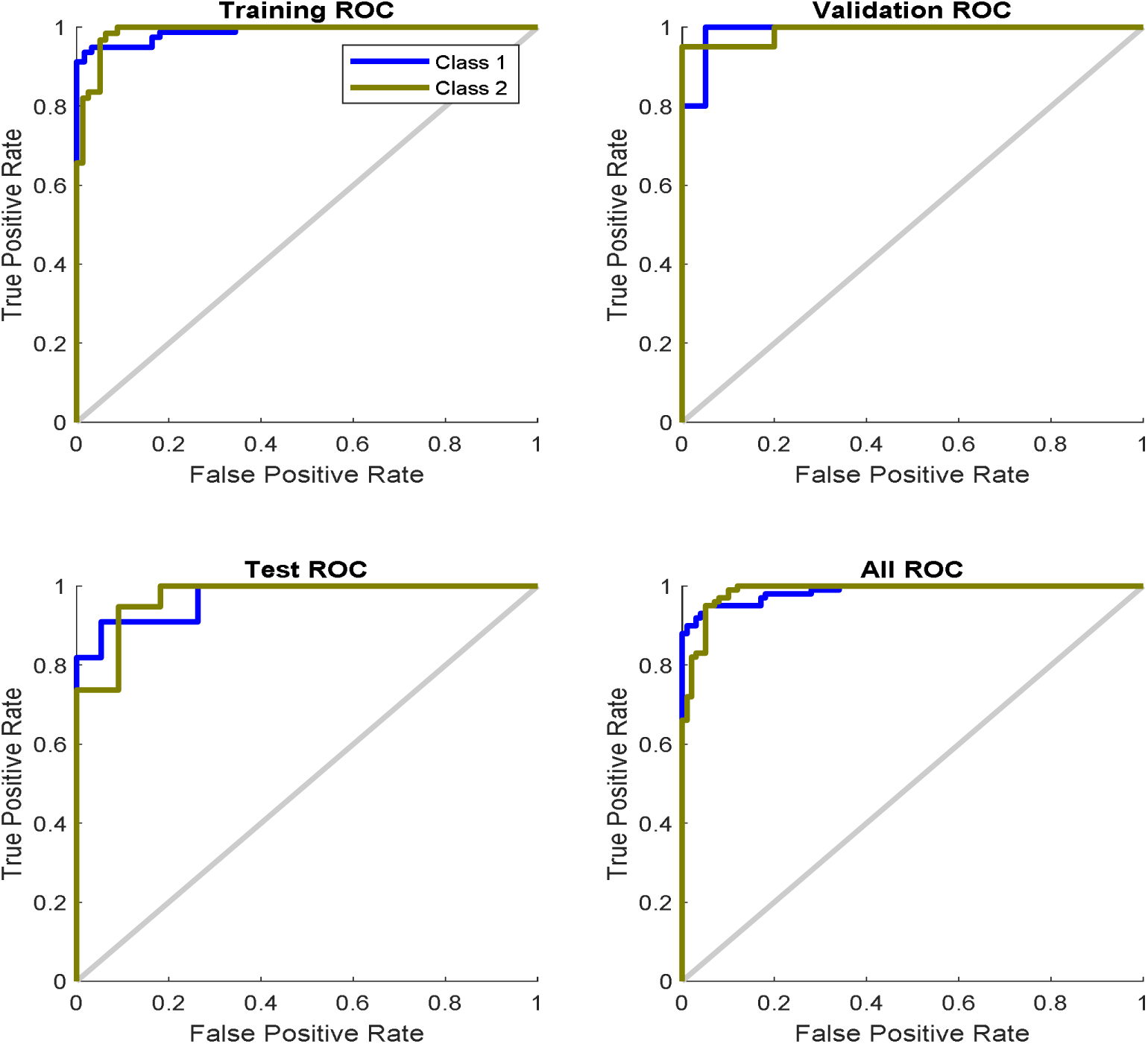
ROC (Receiver Operator Characteristic) curves

**FIGURE. 4.**
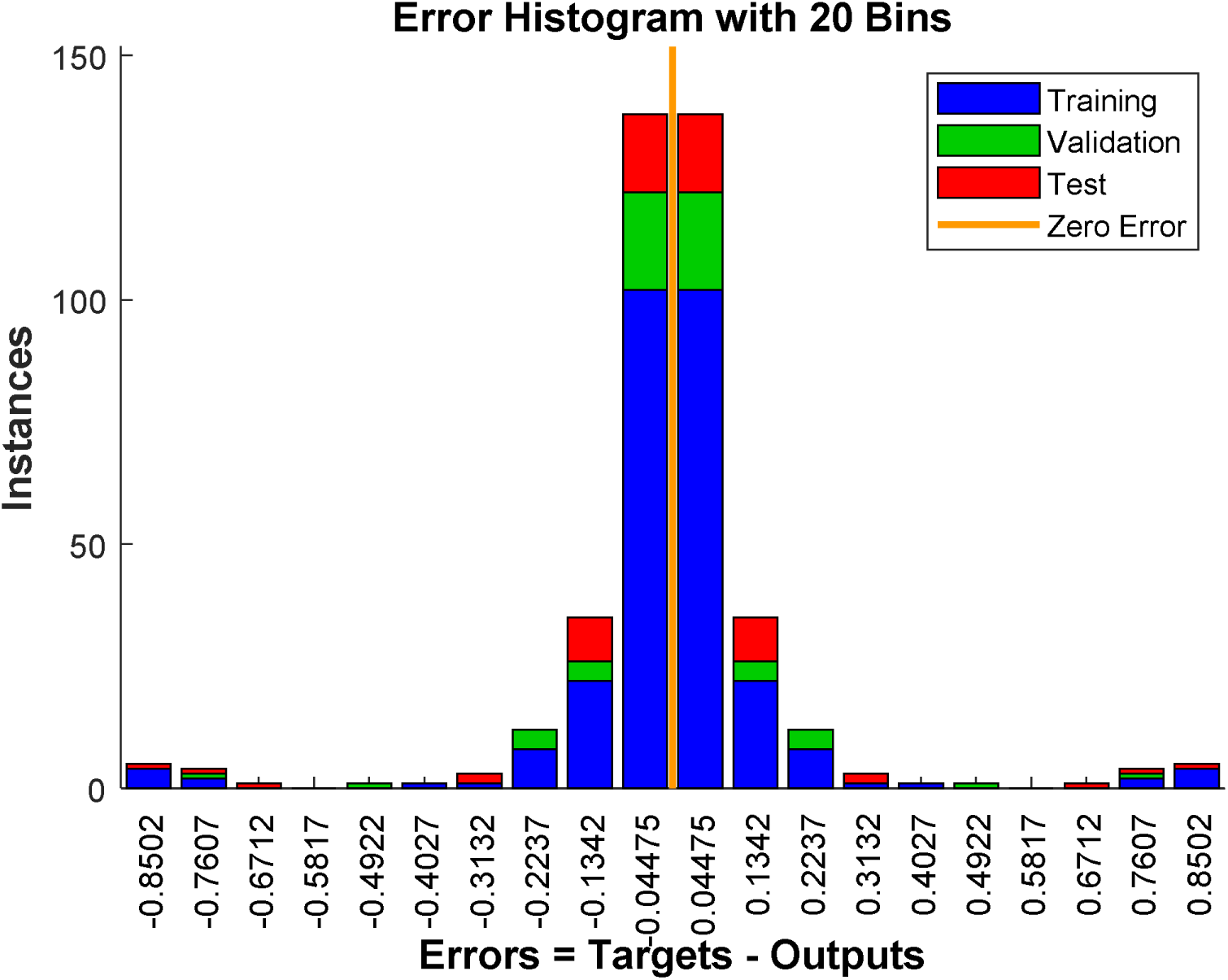
Error Histogram

## DISCUSSION

Artificial Intelligence aided analysis of images to differentiate lesions is an area of interest in the current era. The implementation of neural networks in differentiating malignant and benign breast lesions using grayscale intensity histogram is yet to be explored. In this study, we proved the significance of grayscale intensity histogram and its usage in neural networks to classify benign and malignant lesions.

We have analyzed only one ultrasound parameter of the lesions and the conclusions were derived. The ROIs were drawn manually in this study which is time-consuming and can result in inter-observer variability. The above-mentioned limitations can be corrected by including multiple imaging variables that correlate with clinical findings and by using automated systems respectively.

The positive predictive value of the CNN was 95%. This suggests that the Convolutional Neural Network (CNN) analysis of the grayscale intensity histogram of a USG image of breast lesion is an easy and effective method to differentiate between malignant and benign nature of the lesion. Further studies are needed to increase the robustness of the findings of our study.

## Data Availability

The datasets analyzed during the current study are available in the https://data.mendeley.com/datasets/wmy84gzngw/1

## Notes

### Competing Interest Statement

The authors have declared no competing interest.

### Funding Statement

None. No funding to declare.

